# Pharmacogenomics in Tailoring First-Line Therapy for Breast Cancer: PharmGKB Database Insights

**DOI:** 10.1101/2023.11.28.23299155

**Authors:** Murilo Porfírio de Aguiar

## Abstract

This study delves into the role of pharmacogenomics in optimizing first-line breast cancer treatments, leveraging insights from the PharmGKB database. It emphasizes the importance of understanding genetic variations and their influence on the efficacy and side effects of key drugs like Letrozole, Anastrozole, Tamoxifen, Trastuzumab, and Pertuzumab. By analyzing these drugs in the context of genetic profiles, the research highlights the necessity of incorporating genetic testing into treatment planning. The findings underscore the potential of pharmacogenetics to personalize therapy, aiming to improve outcomes and reduce adverse reactions in breast cancer patients. This approach marks a significant stride towards tailored cancer treatments, underscoring the need for continued research in this field.

## INTRODUCTION

Breast cancer is a major global health concern, with age-standardized incidence and mortality rates estimated at 37.4 and 13.2 per 100,000 women, respectively [1]. Recent advances in personalized medicine have significantly increased interest in pharmacogenetics, a field that examines how genetic variations influence an individual’s response to medications [2, 3]. This area is particularly critical in breast cancer treatment, where the effectiveness of therapy is often undermined by drug resistance and treatment-related toxicities [1].

The goal of pharmacogenetics is to identify individuals who are at an increased risk of experiencing toxicity at conventional doses of chemotherapeutic agents. Genetic variations in genes responsible for drug metabolism, transport, or targeting are known to contribute to the interindividual variability observed in both the efficacy and toxicity of drug treatments [4]. Of particular note are the polymorphisms in the CYP2D6 gene, which have been linked to the metabolism and clinical efficacy of tamoxifen, a cornerstone in the pharmacological management of breast cancer [1].

In the realm of first-line breast cancer treatments, drugs such as letrozole, anastrozole, tamoxifen, trastuzumab, and pertuzumab are commonly employed. These agents have demonstrated substantial effectiveness; for instance, letrozole has been shown to offer superior quality-adjusted survival rates compared to tamoxifen in a large-scale study involving over 900 patients with metastatic breast cancer [5]. The inclusion of these drugs in this integrative review reflects their widespread use and proven effectiveness in managing breast cancer.

The PharmGKB database serves as an extensive resource that compiles and curates knowledge on the impact of genetic variations on drug responses. This database gathers, curates, and disseminates information on clinically relevant gene-drug interactions and genotype-phenotype correlations [2, 3]. The application of PharmGKB in this integrative review provides a robust basis for examining how pharmacogenetics is being integrated into the personalization of first-line therapies for breast cancer.

This review endeavors to contribute to the scientific community by offering an analysis of how pharmacogenetics can be utilized to predict patient responses to first-line medications in breast cancer therapy, taking into account genetic variations and their effects on drug metabolism and efficacy. A more comprehensive understanding of these mechanisms can lead to the development of more effective and personalized treatment strategies for breast cancer.

## METHODOLOGY

This study was formulated to answer the question: “How is pharmacogenomics applied to personalize first-line therapies in breast cancer treatment?” We undertook an integrative review of the PharmGKB, an international pharmacogenetics database, focusing on five primary breast cancer drugs: Letrozole, Anastrozole, Tamoxifen, Trastuzumab, and Pertuzumab. These drugs were selected based on their widespread use and demonstrated effectiveness in breast cancer therapy. Each plays a vital role in treatment, and their therapeutic responses may be affected by genetic variances. For each medication, we gathered all relevant “Clinical Annotations,” particularly focusing on the gene, the variant, and the characteristics of carriers of each allele. We then collected “Variant Annotations” for all variants referenced in the clinical citations. Clinical annotations without corresponding variant information were excluded. The evidence level for each variant’s data and the demographic origin of each study were also reviewed.

Through this methodology, we identified 68 clinical annotations related to key first-line drugs for breast cancer in the PharmGKB database. Pertuzumab, a monoclonal antibody used in HER2-Positive breast cancer, was the only drug without clinical data in the database. For each clinical annotation, we verified the presence of “variant annotations,” which detail the genomic location of the variant, how it alters the genetic sequence, and its potential impact on gene function. Nine clinical annotations lacking such details were omitted. Additionally, 3 clinical annotations were removed due to data redundancy, resulting in a total of 12 clinical annotations being excluded and 56 deemed eligible in the selection and eligibility phase. During the inclusion phase, each eligible clinical annotation was rigorously analyzed to rule out data without sufficient statistical support or consideration of ethnic genetic differences. Consequently, 44 clinical annotations were further excluded, leaving 12 for inclusion in this review. Notably, none of the studies involving Trastuzumab met our criteria, as illustrated in:

**Figure 1.**
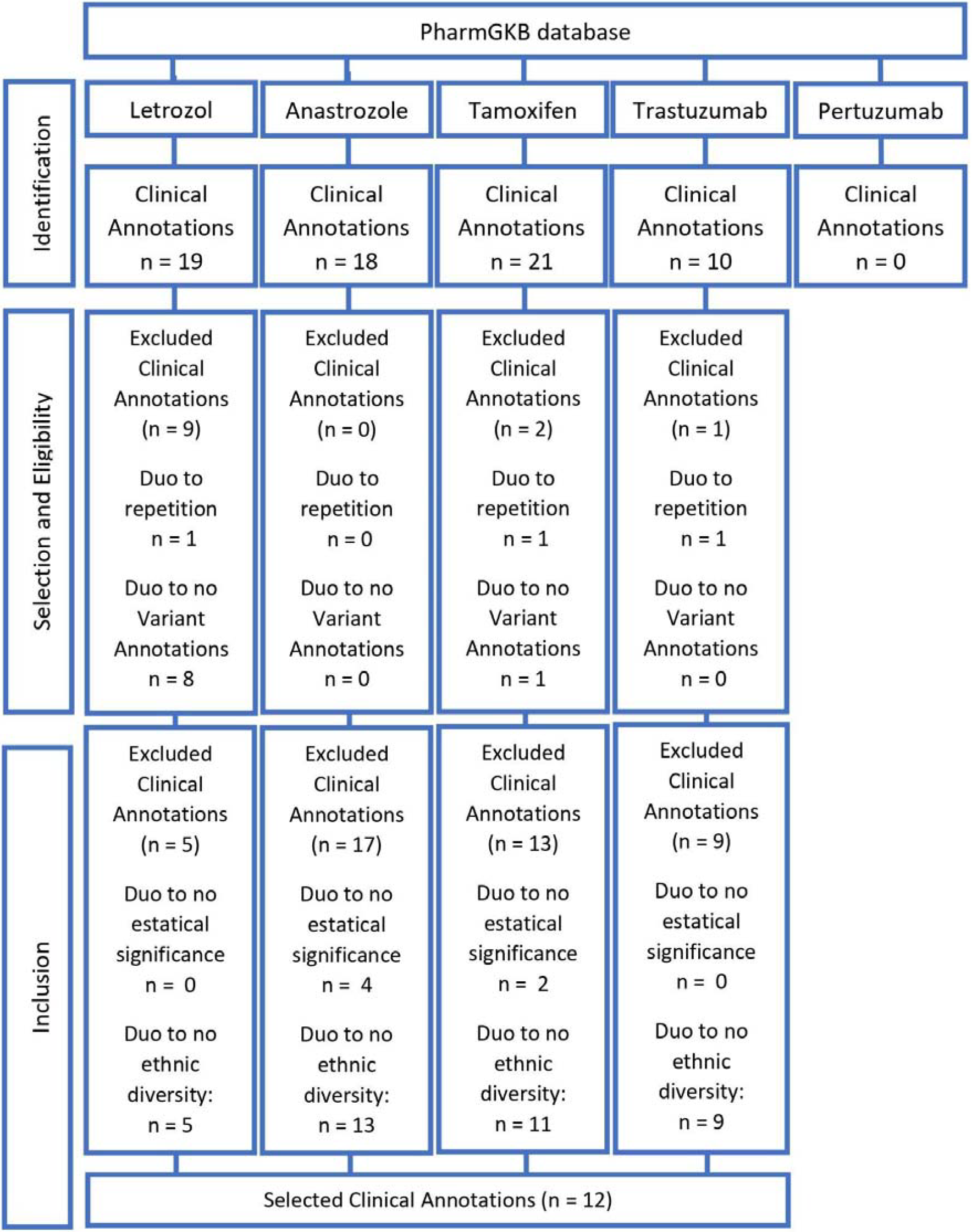
Flowchart of Study Search and Selection.

## RESULTS

The integrative review on the application of pharmacogenetics in the treatment of breast cancer with first-line drugs has revealed significant findings. It has been observed that specific genetic variations in patients impact the efficacy and side effects of the drugs Letrozole, Anastrozole, and Tamoxifen.

For Letrozole, genotypes in various genes such as CYP19A1, CYP2A6, ESR1, and ESR2 have been identified as influencing the risk of bone density loss and the drug’s metabolization rate. This information is crucial for dose adjustment and predicting treatment response in breast cancer patients.

In the case of Anastrozole, the UGT1A4 rs3732218 genetic variant has been shown to affect the drug’s glucuronidation. This indicates variability in the drug’s elimination from the body, which can influence both its efficacy and the incidence of side effects.

Regarding Tamoxifen, variations in the CYP2D6, ABCC2, ESR1, ESR2 genes, and a non-coding region have been demonstrated to significantly impact cancer recurrence, bone fracture risks, and alterations in cholesterol and triglyceride levels. These findings underscore the importance of considering the genetic profile in the selection and monitoring of Tamoxifen treatment.

**Table 1.** Pharmacogenetic Correlations in Breast Cancer Treatment.

| Drug | Gene / Variant | Annotation description | Authors |
| --- | --- | --- | --- |
| Letrozol | CYP19A1 / rs6493497 | In breast cancer treatment, women carrying the AA genotype may experience a heightened risk of bone density loss when treated with exemestane and letrozole. This risk contrasts with those possessing AG and GG genotypes, who typically exhibit a reduced risk. | Liewei et al. [6] |
|  | CYP2A6 / CYP2A6*1 | Patients with dual copies of the CYP2A6*1 allele or a combination of *1 and *46 alleles tend to metabolize letrozole at an accelerated rate compared to individuals with other distinct genetic combinations. This rapid metabolism stands in contrast to that observed in patients with allele combinations including *2, *4, *7, *9, *12, *17, among others. | Desta Z et al. [7] |
|  | ESR1 / rs9322335 | Women with breast cancer and the CC or CT genotypes are likely to have a diminished risk of bone mineral density loss when undergoing treatment with letrozole and/or exemestane, unlike those with the TT genotype. | Steffi et al. [8] |
|  | ESR1 / rs4870061 | In breast cancer patients, those with CC or CT genotypes may experience a lower rate of bone density loss when treated with letrozole, whether or not combined with exemestane, in contrast to individuals with the TT genotype. | Steffi et al. [8] |
|  | ESR2 / rs10140457 | Women diagnosed with breast cancer who have the AC genotype might face a reduced risk of bone density loss when treated with letrozole, unlike those carrying the CC genotype. It's noteworthy that women with the AA genotype were not included in this particular study. | Steffi et al. [8] |
| Anastrozole | UGT1A4 / rs3732218 | Patients carrying the AA or AG genotypes are likely to show decreased glucuronidation of anastrozole compared to those with the GG genotype, as demonstrated by in vitro studies. Efficient glucuronidation is vital for the elimination of xenobiotics, including anastrozole. | Koroth et al. [9] |
| Tamoxifen | CYP2D6 / CYP2D6*1 | The presence of the CYP2D61 allele, identified as a normal-function allele, is linked to a decreased likelihood of recurrence and improved event-free survival in patients undergoing tamoxifen treatment for breast cancer. This association is particularly notable when the CYP2D61 allele is combined with other normal metabolizer alleles, as opposed to combinations including null or reduced function alleles. | Goetz et al. [10] |
|  | ABCC2 / rs717620 | In breast cancer patients positive for ER and/or PR, those with the CC genotype may experience a lower incidence of bone fractures when treated with the aromatase inhibitors anastrozole and exemestane, compared to those with CT or TT genotypes. Patients with the CT genotype face a higher fracture risk than those with CC but lower than those with TT. The TT genotype is associated with the highest fracture risk in these circumstances. | Insee et al. [11] |
|  | ESR1 / rs9340799 | Women carrying the AA genotype exhibit a less pronounced reduction in total cholesterol due to tamoxifen treatment in postmenopausal women and minor variations in triglycerides and HDL in premenopausal women, compared to those with the GG genotype. Those with the AG genotype display intermediate effects on postmenopausal total cholesterol and on triglycerides and HDL levels in premenopausal women, in comparison to the GG genotype. Individuals with the GG genotype experience a more significant decrease in total cholesterol in postmenopausal women and greater fluctuations in triglycerides and HDL in premenopausal women, in contrast to GA/AA genotypes. | Ntukidem et al. [12] |
|  | ESR2 / rs4986938 | In postmenopausal individuals, those possessing the CC genotype are likely to exhibit a notably higher increase in triglyceride levels as a response to tamoxifen treatment, in contrast to those with TT or CT genotypes. | Ntukidem et al. [12] |
|  | ZNF423 / rs8060157 | Individuals carrying either the AA or AG genotypes might face a diminished risk of breast cancer onset while undergoing treatment with Selective Estrogen Receptor Modulators (SERMs), such as raloxifene or tamoxifen, relative to those with the GG genotype. | James et al. [13] |
|  | Non-coding DNA / rs10030044 | Patients with GG or GT genotypes have an increased risk of breast cancer when treated with Selective Estrogen Receptor Modulators (SERMs) like raloxifene or tamoxifen in breast neoplasms, compared to those with the TT genotype. Conversely, TT genotype carriers may have a decreased risk compared to GG genotype individuals. | James et al. [13] |

## DISCUSSION

In examining the role of pharmacogenetics in breast cancer treatment, it’s essential to recognize the limitations of current genetic panels: they often lag behind the latest research developments and tend to focus on a narrow range of genetic variations. This approach can overlook critical genetic variations that have emerged as statistically significant, while still including those that have become less relevant [1]. Additionally, these studies predominantly target individuals of European descent, resulting in a lack of genetic diversity. This narrow focus can lead to research outcomes that do not adequately represent the entire global population, potentially limiting their applicability across various ethnic groups or individuals with diverse genetic backgrounds [14].

Another significant hurdle in the clinical adoption of pharmacogenetics is the associated high cost. Equipping and maintaining laboratories with the latest technology and skilled personnel to perform and interpret genetic testing, or outsourcing these services, presents a substantial financial challenge, especially for healthcare facilities in developing countries [15].

Despite its vast potential, pharmacogenetics encounters substantial obstacles, including the need for frequent updates to genetic panels to encompass a wider array of genes and the challenge of managing the high costs associated with this advanced technology and specialized training. Addressing these challenges is imperative for optimizing the use of personalized medicine in breast cancer treatment, ensuring it benefits patients from all ethnic and socioeconomic backgrounds.

## CONCLUSION

Pharmacogenomics is applied to personalize first-line breast cancer treatments through the identification of genetic variations that impact the efficacy and side effects of drugs like Letrozole, Anastrozole, and Tamoxifen. This approach involves analyzing specific genotypes, such as CYP19A1, CYP2A6, ESR1, ESR2, and UGT1A4 rs3732218, to understand their influence on drug metabolism and patient response. This methodology enables more accurate dose adjustments and treatment plans tailored to individual genetic profiles, thereby enhancing treatment effectiveness and reducing the risk of adverse effects. The study highlights the critical role of pharmacogenetics in advancing personalized medicine in breast cancer treatment, emphasizing the need for further research to explore the impact of various genetic variations on drug response.

## Data Availability

All data produced in the present work are contained in the manuscript.

